# Physiological and Psychological Effects of Parental Traditional Thai Massage on Children with Autism: A Protocol for a Randomized Controlled Trial

**DOI:** 10.1101/2021.12.22.21268297

**Authors:** Hui Ruan, Wichai Eungpinichpong, Hua Wu, Chanada Aonsri

## Abstract

**Introduction:** Although many autistic children receive massage as a complementary therapy, it is not included in evidence-based practices for autism since evidence of its efficacy is lacking. Further, prior studies have failed to identify objective indicators of core symptoms or elucidate their mechanisms. We developed a parent-delivered traditional Thai massage (TTM) on children intervention with autism, aim is to access the physiological (gait and heart rate variability) and psychological effects of intervention.

**Methods and analysis:** A two-armed, parallel randomized controlled trial study will recruit forty-eight children with autism from the XX Special Education School at the beginning of Febuary2022. They will be randomly assigned to either a parental TTM or control group with a ratio of 1:1. Individuals in the parental TTM group will receive 16 parent-delivered TTM sessions over 8 weeks. The outcome will be assessed on admission, after 8 weeks, and as well as at 2-month follow-up, including the Autism Treatment Evaluation Checklist score, heart rate variability, gait, and parenting stress index.

**Ethics and dissemination:** Ethical approved by the Khon Kaen University Ethics Committee for Human Research (XXXX). Result will be published in a peer-reviewed journal and give presentations in domestic and international academic conferences to further promote communication.

**Trial registration number:** XXXXXX

**Strengths and limitations of this study:**

1. This will be the first RCT to explore parental TTM in terms of its physiological and psychological effects on children with autism.
2. Compared with previous studies, we access the objective index to discuss the possible mechanisms of observed effects.
3. Our study may highlight the value of complementary and alternative therapies for adjunctive treatment of autism to enhance family-based interventions
4. Both parents and blind teachers will evaluate for effects to decreases the subjectivity of bias. And the variability massage manipulation by parents is minimized by training with experts, examinations, and supervision.

## 1 INTRODUCTION

Autism is a neurodevelopmental disorder that affects central nervous system development, and core symptoms is characterized by social interaction and social communication deficit, and restrictive repetitive behavior patterns[1]. The disorder is not associated with any particular culture, ethnicity, race, or socioeconomic group[2]. The majority of autism still have motor disorders, sensory disorders, anxiety, hyperactivity, sleep disorders, and autonomic nervous system (ANS)-associated disorders[3–5]. The prevalence of autism has risen rapidly throughout the last decade. An assessment commissioned by the WHO in 2012 estimated the global prevalence of autism be approximately 1%[6]. The updated report from CDC showed one in 44 children aged 8 years was estimated to have ASD recently[7]. The survey found the incidence of autism among children in China is comparable to that in Western countries[8].

The causes of autism are largely unknown and medically difficult to cure, which has promoted the development a variety of non-pharmacological therapies. Although effects of complementary and alternative medicine are uncertain[9], its accessibility, relatively low cost, and lack of substantial side effects are attractive characteristics noted by families of autistic patients [10].Massage is often used as a complementary therapy in the treatment of autism. It has been reported that 11%–16% of people with autism undergo massage therapy[11].

Abnormalities associated with touch most greatly impacted social delays in individuals with autism. A theoretical sensory processing model proposed by Dunn considers attention, arousal, emotion, and activity[12],which provides guidance for interventions that aim to improve sensory impairment in autism and may significantly impact core symptoms. Massage is a touch-based therapy[13] in which patients undergo force mechanical stimulation. Force provided by massage provides suitable stimulation to a tactile sensor. Previous studies have shown that massage improves social communication[14,15], stereotypic behavior [16,17], sensory profiles [18,19], language ability [20,21] in autism. Traditional Thai massage (TTM) has been used for thousands of years, with its three distinctive signatures: deep pressure massage, work on meridian (energy) lines, and combine with stretching to the affected muscles, and joints. TTM are widely used in health care[22,23], rehabilitation[24,25],and so on. Evidence has shown that TTM may facilitate immediate and short-term improvement of heart rate variability (HRV) [26,27] and some gait parameters in patients[28,29].It has also been used effectively in the rehabilitation of children with disabilities by the Bangkok Foundation for Children with Disabilities[30]. However, the existing massage research on autism mainly came from effects of qigong massage and Tui na intervention measured by scales, only one study[31] has assessed the use of TTM to improve symptoms of autism, and the effect of TTM in children with autism remains unknown.

The Lancet Commission latest findings highlight the use of resources to support autism care and improve the quality of life for families with autism[32]. Previous research revealed that parents who engaged in the intervention not only satisfied family based care needs[33],but also strengthened the parent-child bond[18,19,34].Our two-armed with a ratio of 1:1, parallel, exploratory randomized controlled trial aims to assess physiological and psychological effects of parent-delivered TTM in children with autism and determine the possible mechanisms of observed effects.

## 2 METHODS: STUDY DESIGN, PATIENTS, AND INTERVENTIONS

### 2.1 Objectives of the study

This study aims to assess the effects of parental TTM in children with autism by evaluating physiological (gait and HRV) and psychological parameters (autism symptoms questionnaires). The primary objective of the study is to assess the effectiveness of parental TTM in improving symptoms of autism by scale. Secondary objectives are to evaluate instant and cumulative effects of the parental TTM intervention via an assessment of gait and HRV, to explore mechanisms of symptom improvement and observed physiological parameter changes, to examine changes in levels of parental stress, and to observe differences in relationships with children after parents administer the TTM intervention.

### 2.2 Research questions

The following four research questions will be addressed:

1. Do core symptoms improve in autistic children after parental TTM in comparison with the control group?
2. Do HRV and gait indicators improve in children with autism after parental TTM?
3. Do core symptoms change in association with the improvement of physiological indicators?
4. How do parent-child relationships change throughout the intervention period measured by Parenting Stress Index?

### 2.3 Design and setting

A randomized controlled trial (RCT) will be conducted. Participants will be recruited from XX Special Education School, XX Province, XX. Recruiting poster and presentation will be post it on the bulletin board. In accordance with SPIRIT guidelines[35](Supplementary File 1), a flowchart of the study protocol is depicted in Figure 1. Before recruiting participants, informed consent will be sent to parents via the aid of the teacher, and the parents will submit to the designated box in two days.

**Figure 1.**
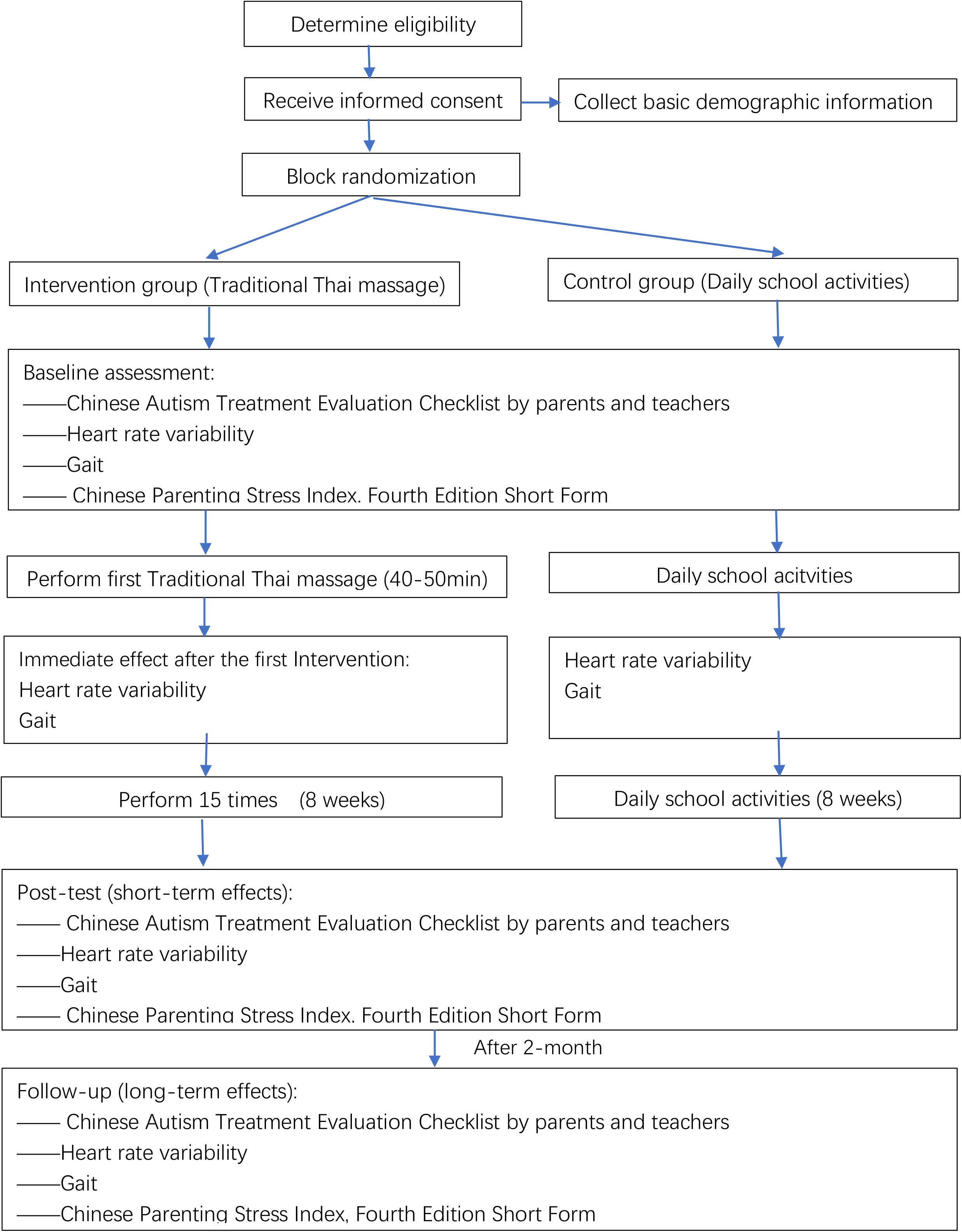
Flowchart depicting the study design.

### 2.4 Participants

#### 2.41 Inclusion criteria

The following inclusion criteria will be applied to select among autistic child: diagnosed with autism by a child psychologist, aged 7 to 12 years, not receiving systemic massage therapy, and able to follow instructions. Children`s primary caregivers must meet the following criteria: have been providing ≥1 year of child care; able to communicate with massage trainer. Teachers must meet the following inclusion criteria: serve as primary caregivers for autistic children at school; provide care for at least one semester and be willing to participate in research.

#### 2.4.2 Exclusion criteria

For autistic child, the following exclusion criteria will be applied: contraindications of TTM, such as fractures, fever, psychoactive medication; and administration of other specialized interventions during the experimental period. The exclusion criteria for parent are failure to attend all required TTM theory and practice. Further, teachers with insufficient time to complete the assessment will be excluded.

#### 2.4.3 Sample size

The sample size calculation used is based on Najafabadi et al research[36], due to no study used both the autism massage intervention and the ATEC scale. The standard deviation of social ATEC scores of the two groups will be used to calculate the sample size needed to achieve a power of 80% and a significance level of 5%. A dropout rate of 10% will be assumed to determine the principal of intention for estimating the final sample size needed. According to the formula[37], a total of 48 participants will be recruited for the study.

#### 2.4.4 Randomization

Forty-eight participants (7 –14 years of age) from 1to 6 grade will be randomly assigned to parental TTM and control groups by random numbers through Online Research Randomizer software. The sequences generated will then be concealed in an opaque envelope.

#### 2.4.5 Blinding

Teachers will be blinded to the group assignments of children included in the study. Parents will be asked to refrain from discussing the intervention, which prevents the contamination of the control group. Data collectors will also be unaware of group placement. Data will be renamed and converted by the second author.

### 2.5 Intervention

Children assigned to the TTM group will receive sixteen 40–50-minute parent-delivered TTM sessions over an 8-week period (two sessions per week) after class in the health room of school. Gifts and rewards will give to parents and children to increase intervention adherence rates. The control group will maintain a normal daily routine when in school.

#### 2.5.1 TTM training

Parent of autistic children who are enrolled in the study will take part in three TTM training sessions. Each 3-hours session will be hosted by experts at the Indoor Activity Center of XX Special Education School. Parents of participants in the intervention group will be trained in TTM by an experienced traditional Thai masseur with a massage qualification, until becoming eligible to perform the experimental intervention. Thereafter, massage trainer will monitor the parents to ensure that they continue to perform TTM properly at each session.

#### 2.5.2 TTM procedure

The TTM method involves pressure massage and muscle stretch: 1) pressure massage: massaging with thumbs and palms press until slight muscle resistance is felt. The children may feel slight muscle tightness but the massage should not be painful. The masseur will maintain pressure for 5–10 seconds, and then release it thereafter to allow energy or blood to flow freely around the massage area, repeat 2-3 times for each body part. 2)muscle stretch: when performing stretches, the masseur asks/observes the children about the extent to which he/she feels the stretch to determine that the stretch is not too deep, and holds the appropriate position for 15 to 30 seconds[38].

In the starting position, the children lie on his/her side with the upper leg bent at the knee forming a 90° angle, pillows placed under the head, and a straight lower leg. The person performing the massage will kneel or sit behind the child. Massage strategies for distinct massage types should be performed in accordance the following specifications (Figure 2).

**Figure 2.**
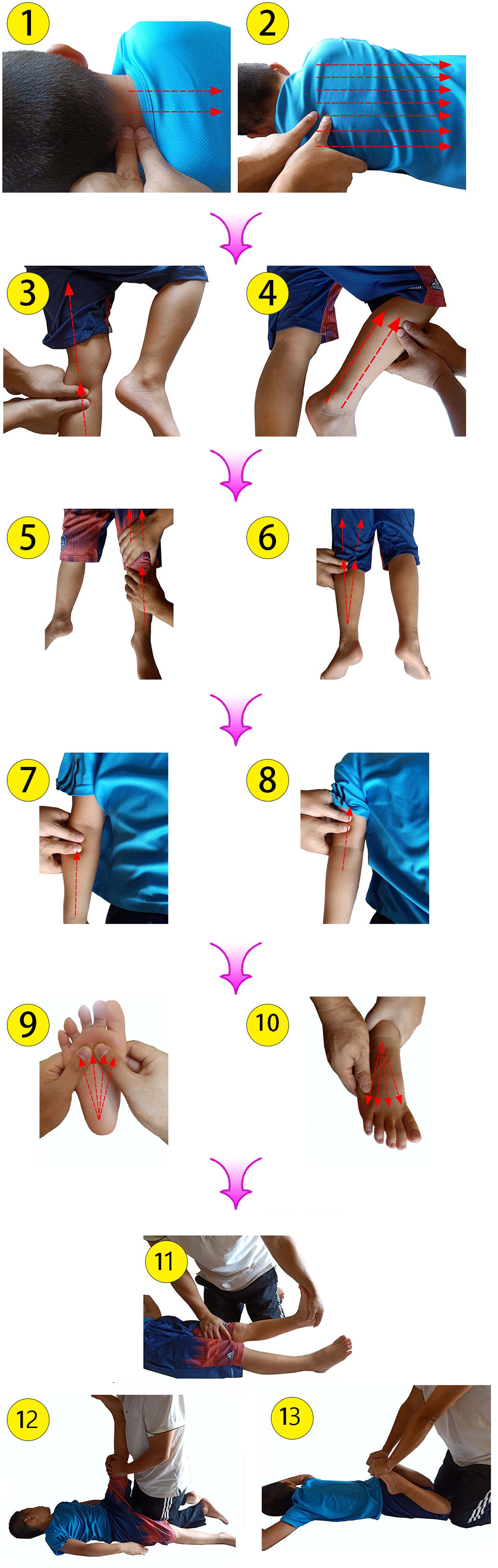
Traditional Thai massage procedure ① Neck massage; ②Back and waist massage; ③ Inner thigh massage; ④ Outer leg massage; ⑤ Leg massage in the supine position; ⑥Leg massage in the prone position; ⑦⑧Arm massage; ⑨⑩Foot message; (11-13) Muscle stretch.

1. Neck massage: First, the sides of cervical muscles should be massaged, starting in the lumbar region, and extending along back muscles toward the lower cervical area (C7). Next, the masseur should massage along the side of the neck toward the shoulders.
2. Back and waist massage: The paravertebral muscles, which are close to the thoracolumbar spine, should be massaged toward the waist level. There are 3 lines of massage. One is started on the upper back near the spinous processes of T1 to L5 on each side of the back. The second line is about 1 centimeter along the side of the first one. The third line is about 1 centimeter along the side of the second one. The pressing massage style should be applied on the three lines, which should be initiated in the thoracolumbar region and continue toward the top of the sacrum.
3. Inner thigh massage (straightened legs): When performing inner thigh massage, thumb pressure should be applied to the medial side of a straightened leg from the Achilles tendon toward the groin (line #3 in the picture). This type of massage should be performed along two message lines. The first massage line extends along the medial side of the leg, while the second massage line extends along the backside of the leg.
4. Outer leg massage (on the bent leg side): The masseur should place both hands on the lower leg of the child. The lateral part of the lower leg should be pushed down using the thumb. The massage style should be applied from the area above the lateral malleolus and continued to the side of the knee joint. Next, the masseur should press from the top of both knees toward the middle of the hip. Outer leg massage should be performed along the two massage lines. The first line extends along the front of the leg, and the second line is positioned slightly behind the first line. Opposite side massage: The masseur asks the volunteer to lie on the other side, then he repeats massage procedures 1–4, as described previously.
5. Leg massage in the supine position: The child lies on his/her back, the masseur begin the massage in front of ankle to near the groin.
6. Leg massage in the prone position: With the children on his/her stomach, the masseur initiates the massage behind the heel around the Achilles tendon and continues toward gluteal and hamstring muscles.
7. Arm massage: With the children lying on his/her stomach, the masseur should provide thumb pressure along the back of the lower and upper arm toward the shoulder. The procedure is then repeated on the other arm of the children. Next, the children should lie on his/her back, with arms extended in a supine position, and the masseur facing their head. In this position, the masseur should push upward from the center of the forearm, and along the center of the arm toward the axilla.
8. Foot message: With the child children lying on his/her back, the masseur performs a foot massage by pressing from heel to all five toes. Then, the tip of each toe should be pulled gently.
9. Muscle stretch: The purpose of stretching the muscles is to allow child to feel maximum muscle tension without pain. When performing stretches, the masseur asks the children about the extent to which he/she feels the stretch to determine that the stretch is not too deep, and holds the appropriate position for 15 to 30 seconds. The following stretches should be performed:

1. The child lies on his/her back while the masseur stretches the heel,
2. With the child lying on his/her back and the masseur standing, the masseur grabs the volunteer’s foot, lifting it until tension is felt.
3. With the child lying on his/her stomach, the masseur flexes the child volunteer’s knees as much as possible by applying gentle pressure to the soles of his/her feet.

## 3 METHODS: OUTCOME ASSESSMENT

### 3.1 Primary outcome measurement

#### 3.1.1 ATEC

The ATEC questionnaire is developed by Dr. Bernard Rimland and Dr. Stephen M. Edelson from the Autism Research Institute of USA in the mid-1990s, which designed to help researchers or parents evaluate the effectiveness of various treatments for autistic children and adults. It is comprised of four subscales: speech/language/communication, sociability, sensory/cognitive awareness, and health/physical/behavior. The maximum score is 179, with higher scores indicating autism of increased severity. The sensitivity and specificity of the Chinese version of the ATEC total scale and its subscales were 0.922∼0.987 and 0.803∼0.887, respectively, and the scale has been determined have high reliability[39].

### 3. 2 Secondary outcome assessment

#### 3.2.1 Gait

Gait will be measured using a wearable BTS G-Walk device (BTS Bioengineering Company, Lombardia, Italy). The experimental gait data will be collected at a frequency of 100 Hz, and transmitted to a computer via a Bluetooth connection. All gait parameter measurements have been determined to have high test-retest and inter-trial reliability (ICC: 0.728-0.969) (ICC: 0.84 -0.99)[40,41]. Wireless internet is required for gait tests, which must be carried out on a flat surface that measures at least 10 m × 10 m. Prior to performing the test, the height of a participant will be entered, and an adjustable elastic band should be worn around the participant’s waist. When taking the test, participants will stand in place for a few seconds. After confirming that the sensor is connected to the computer via Bluetooth and the connection is stable, each child volunteer will continue following a marked, 7-meter path, back and forth, while walking normally. Teachers or parents may provide verbal cues but will not be permitted to provide physical assistance.

#### 3.2.2 HRV

UBiomacpa V1 (Biosense Creative, Korea) will be used to detect the activity of the autonomic nervous system. The HRV test will be carried out in an independent and quiet room at an ambient temperature of 26°C. Before starting the test, the tester should ensure that equipment is properly connected. After receiving TTM, autistic child will be placed in a sitting position with a pulse sensor attached to their left index finger and undergo an HRV test for 2 minutes that is administered by a physiotherapist. When the test is complete, the device will automatically generate a data report via a connected computer.

#### 3.2.3 Parenting Stress Index, Fourth Edition Short Form (PSI-4-SF)

The PSI-SF scale consists of 36 items in the following three dimensions: parent distress, parent-child dysfunction interaction, and difficult child [42].The Chinese Version of the PSI-4-SF has an internal consistency and reliability that ranges between 0.92 and 0.95 for each subscale. For the whole scale, the value is 0.97, indicating that this scale displays good consistency and stability[43].

## 4 METHODS: DATA COLLECTION, MANAGEMENT, AND ANALYSIS

IBM SPSS 25 (IBM, Armonk, USA) will be used for data analysis. Missing outcome data will not be imputed in the analysis, but the multiple imputations method to explore the potential impact of missing data on outcomes. Demographic data will be expressed as means ± standard deviation and percentages. The purpose of this is to compare effects of TTM on autistic children at including immediate, short-term, and long-term time-points. Generalized estimating equations will be used to assess the following: time = covariate, model: group, time, group x time. The relationship between physiological effects and ATEC results will be evaluated using a correlation test. A coefficient value of < 0.4 indicates a ‘low’ degree of correlation, a value between 0.4 and 0.7 indicates a ‘medium’ degree correlation, and a value > 0.7 indicates a ‘high’ degree of correlation between variables. No plans for a subgroup analysis currently.

## 5 METHODS: MONITORING

### 5.1 Data monitoring

The exploratory RCT has been designed for simplicity and minimal risk, hence no formal data monitoring committee has been established and no interim analysis of the effects of the intervention is planned.

### 5.2 Harms

No adverse events with Thai massage intervention reported in previous studies. Furthermore, both intervention and control groups, the data collection did not cause physical harm to children’s violence, no psychological, social, financial risks. If the risks that may affect body (pain, contusion), and mind (unsatisfied, discomfort, hate, bored) in someone, we will temporarily suspend or stop this participant and record.

### 5.3 Auditing

No audit has been planned at this time

### 5.4 Confidentiality

The research group will keep the data confidential, and the research group will use the children’s passwords to record data, access data, etc. In this study, only the research group will know the data, and the research group will not state the names of the volunteers under any circumstances if the study results are published in the journal. The researcher (C.A.) will be responsible for double-enter the data electronically. Participants’ personal information will be stored in a specific file with a preset code. Data sharing not applicable to this article as no new data were generated or analyzed. The datasets generated or analyzed after the experiments are available from the corresponding author on reasonable request.

### 5.5 Ancillary and post-trial care

Participants can contact the researchers during the trial or one month after participation ends. Participants with serious adverse events will receive timely and appropriate responses if they occur.

## 6 DISCUSSIONS

In practice, massage has been used to treat autism. However, the treatment has not been included in evidence-based practices for autism due to the lack of long-term, extensive scientific research that supports its use. At present, studies of interventions have mainly focused on qigong massage and Tui na. Furthermore, we found that outcome measures of most prior studies were based on questionnaires and scales evaluated before and after the intervention, and lack of studies that have explored mechanisms that underlie core symptom improvement. Presumably, massage adjusts the body and mind via sensory process[44,45], and neural and hormonal regulation (e.g., changing oxytocin levels), and oxytocin is known to be involved in ANS circuit regulation[46], which affects both emotional and social processes[47].

## Data Availability

Data sharing not applicable to this article as no new data were generated or analyzed

## Acknowledgments

We would like to thank the teachers and staff at the school and the students who participated in this study.

